# The SHOW COVID-19 cohort: methods and rationale for examining the statewide impact of COVID-19 on the social determinants of health

**DOI:** 10.1101/2023.10.17.23297146

**Authors:** Amy A. Schultz, Erin Nelson-Bakkum, Maria Nikodemova, Sarah Luongo, Jodi H Barnet, Matthew C. Walsh, Andrew Bersch, Lisa Cadmus-Bertram, Corinne D. Engelman, Julia Lubsen, Paul Peppard, Ajay Sethi, Kristen MC Malecki

## Abstract

**Background:** National and large city mortality and morbidity data emerged during the early years of the COVID-19 pandemic, yet statewide data to assess the impact COVID-19 had across urban and rural landscapes on subpopulations was lacking. The SHOW COVID-19 cohort was established to provide descriptive and longitudinal data to examine the influence the social determinants of health had on COVID-19 related outcomes.

**Methods:** Participants were recruited from the 5,742 adults in the Survey of the Health of Wisconsin (SHOW) cohort who were all residents of Wisconsin, United States when they joined the cohort between 2008-2019. Online surveys were administered at three timepoints during 2020-2021. Survey topics included COVID-19 exposure, testing and vaccination, COVID-19 impact on economic wellbeing, healthcare access, mental and emotional health, caregiving, diet, lifestyle behaviors, social cohesion, and resilience.

**Results:** A total of 2,304 adults completed at least one COVID-19 online survey, with n=1,090 completing all three survey timepoints. Non-Whites were 2-3 times more likely to report having had COVID-19 compared to Whites, females were more likely than males to experience disruptions in their employment, and those with children in the home were more likely to report moderate to high levels of stress compared to adults without children.

**Conclusion:** Longitudinal, statewide cohorts are important for investigating how the social determinants of health affect people’s lives, health, and well-being during the first years of a pandemic and offer insight into future pandemic preparation. The data are available for researchers and cohort is active for continued and future follow-up.

**Key Messages:**

- Mortality and morbidity data emerged during the early years of the COVID-19 pandemic at the national scale and in large cities, yet comprehensive social, cultural, and economic population-level data at the state level was lacking for identifying sub-population trends.
- COVID-19 disrupted lives and affected people differently based on socio-economic status, demographics, family dynamics, geography, health status, and employment.
- SHOW COVID-19 cohort is a unique non-clinical, non-hospital-based sample with pre-COVID-19 baseline survey data and biospecimen and three waves of COVID-19 data and specimen available to examine effects of COVID-19 on the social determinants of health.

## INTRODUCTION

The coronavirus disease 2019 (COVID-19) spread to a devastating pandemic, with the worldwide death toll exceeding 6 million.^1^ In addition to the clinical disease, the pandemic also impacted numerous social and behavioral aspects of people’s lives. Due to the lack of a vaccine or effective antiviral medications throughout 2020, governments and employers implemented non-pharmaceutical interventions (NPIs) such as stay-at-home orders, closures of schools and non-essential businesses, physical distancing guidelines, and mask mandates to slow the spread and reduce the burden on the hospital systems.^2,3^ The pandemic altered people’s daily lives in a myriad of ways, including their employment, income, socialization, and family dynamic, which in turn impacted their physical, emotional, and mental health.^4–12^ Exposure to the virus, and the degree to which individuals were able to cope with and follow stay-at-home orders, varied.^6,13–15^ The effects of the COVID-19 pandemic, state-level, and local community response, and adoption of NPIs varied across the United States. To understand the pandemic’s social and behavioral impact in Wisconsin, in May 2020, a multidisciplinary team of investigators at the University of Wisconsin-Madison developed and launched the Survey of the Health of Wisconsin (SHOW) COVID-19 Survey (i.e., The COVID-19 Survey).

The COVID-19 Survey was developed to collect data on COVID-19 exposures and symptoms, healthcare access, employment, caregiving, and daily behaviors. The survey was administered at three timepoints over the first 18 months of the pandemic among an existing population-based, longitudinal cohort of adults, forming the SHOW COVID-19 Cohort. The SHOW cohort is unique in that it is a non-hospital-based sample and the only statewide representative cohort in the United States modeled after the National Health and Nutrition Examination Survey (NHANES). It includes residents who may not have health insurance or a primary care provider, and those who may otherwise not be represented in biomedical research.

The goal of the SHOW COVID-19 Cohort was to use baseline and COVID-19 Survey information to examine the impact the social determinants of health had on the short and long-term effects the COVID-19 pandemic had on people’s lives, their health, and their well-being. The purpose of this paper is to describe the theory and methods of all three waves of the COVID-19 Survey, as well as the challenges in conducting the survey, and highlight the strength and uniqueness of the cohort and data, which is available for researchers.

### COVID-19 outbreak in the United States and Wisconsin

The first confirmed COVID-19 cases in the United States were recorded on January 20, 2020, in Washington State.^16^ The first documented case in Wisconsin, a state in the upper Midwest of the U.S., was on March 9, 2020.^17^ The Governor declared COVID-19 as a Health Emergency on March 12^18^ prompting Executive Orders to close kindergarten through twelfth grade (K-12) schools^19^and non-essential businesses.^20^ Statewide “Safer at Home” orders were implemented on March 24, 2020,^21^ and a statewide indoor mask mandate was issued on July 30, 2020.^22^ By mid-January 2021, the number of deaths from COVID-19 in the U.S. surpassed 400,000,^17^ and in Wisconsin had risen to 5,211.^23^ Figure 1A displays the statewide events and administration of the SHOW COVID-19 surveys throughout the first 18 months of the pandemic. Figure 1B depicts the 7-day average of new COVID-19 cases and deaths in Wisconsin. Both daily new COVID-19 cases and deaths peaked in late fall and early winter of 2020 into early 2021, just prior to vaccines being available to high-risk groups (December 18, 2020) and the public (April 5, 2021).^24,25^ By August 1, 2021, more than 3 million eligible Wisconsin residents had received at least one dose of the COVID-19 vaccine.^26^

**Figure 1.**
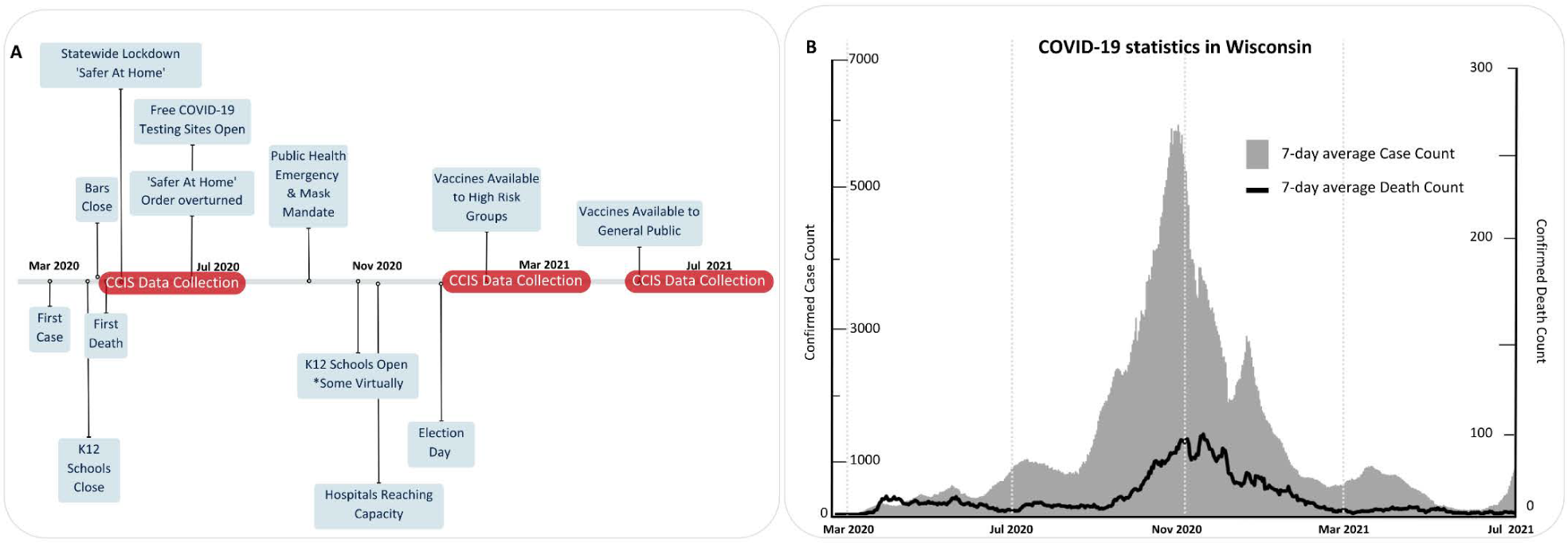
(A) Timeline of COVID-19 events and SHOW COVID-19 Survey waves of data collection and (B) COVID-19 case and death counts in Wisconsin

As the COVID-19 pandemic progressed, disparities in population-level morbidity and mortality emerged throughout the United States.^27^ However, state-level trends in COVID-19 were difficult to ascertain as many state and local health departments lacked infrastructure for tracking beyond the case and death ascertainment. Wisconsin is a geographically and economically diverse state with large metropolitan areas as well as an expansive rural population in both agricultural and forested landscapes. The statewide population-based SHOW cohort provides a unique opportunity to better understand the role the social determinants of health play in the health and well-being of individuals during and after a pandemic.

## METHODS

### Participant recruitment

Participants of the SHOW COVID-19 cohort were recruited from the Survey of the Health of Wisconsin (SHOW) cohort.^28^ The SHOW began in 2008 and includes n=5,742 adults (18-94 years old) that encompass annual and tri-annual statewide representative samples of adults based on probability sampling, as well as an oversampling from traditionally underrepresented populations in biomedical research, including Black/African Americans and Hispanic/Latinos in Milwaukee, Wisconsin. SHOW gathers survey data on health outcomes, and numerous determinants of health including social, economic, current health status, mental wellbeing, diet, and lifestyle factors, as well as objective physical measurements and biospecimen. Details on the SHOW sampling frame, recruitment, and methods are described elsewhere.^29^

Individuals (18+ years of age) of the SHOW cohort who met the following criteria were eligible to be recruited for the COVID-19 Survey: (1) consented to be contacted for future research, (2) have not rescinded their consent to be contacted for future studies, (3) were not deceased as of the first day of recruitment, and (4) had valid email address and/or phone number. Figure 2 depicts a flow chart of the study sample for the COVID-19 Survey. Among the n=5,742 adults in the SHOW cohort, n=767 SHOW adults were ineligible, resulting in n=4,975 eligible adults for recruitment. Eligible adults were invited to participate in all three waves. Participation in one wave had no bearing on being eligibility for future waves. Eligibility was the same for Wave I and II but changed for Wave III. Participants who were minors at the time of their baseline SHOW survey but were 18 years of age on June 1, 2021 (n=215) were additionally eligible for Wave III, increasing the total eligible sample to n=5,190.

**Figure 2.**
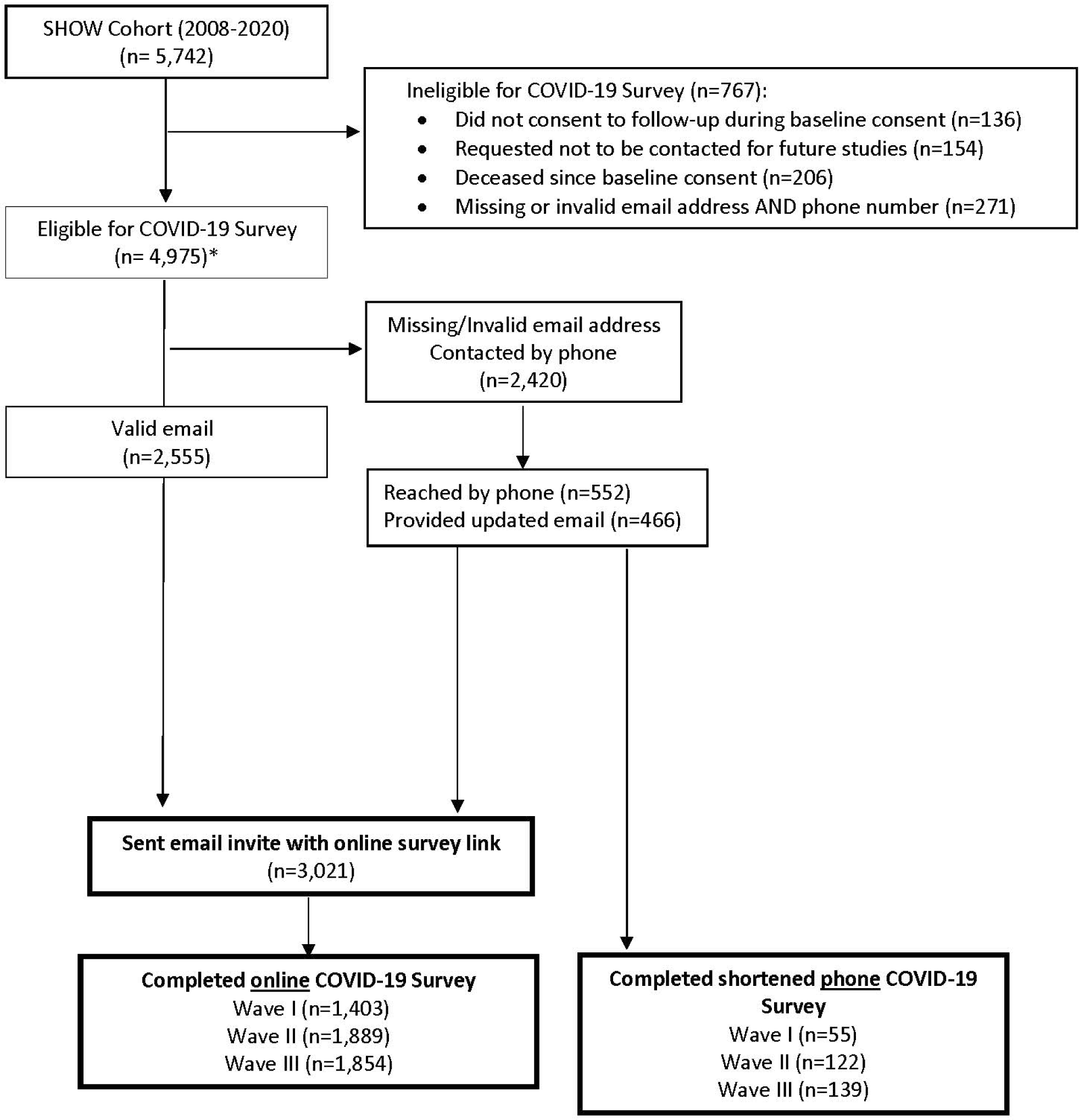
Flow chart of the study sample, depicting eligibility, exclusion criteria, and participation in the SHOW COVID-19 Survey 2020-2021. *For Wave III, n=215 who were minors at baseline but 18 years old by June 21, 2021 were eligible, increasing the total eligible sample to n=5,190 and n=3,236 who were emailed the invite.

As the first COVID-19 survey wave launched, COVID-19 lockdowns and safety precautions precluded SHOW study staff from sending mailed invite letters and surveys and recruiting in-person at their home. Consequently, eligible individuals were emailed invites with a unique online survey link. Those without a valid email address (n=2,420) were contacted by phone. An additional n=466 were sent the online survey link after acquiring a valid email address over the phone, which totaled n=3,021 adults who were recruited via email invite with the online survey link (n=3,236 for Wave III). A tailored, shorter phone survey was made available upon request for participants with limited internet access or literacy.

### The SHOW COVID-19 Survey

The COVID-19 Survey consists of 9 domains including (1) COVID-19 perceptions, beliefs, behaviors; (2) economic well-being; (3) food security, diet, and housing; (4) personal, social and community context; (5) health and healthcare access; (6) mental and emotional health; (7) information sources and literacy; (8) lifestyle behaviors; and (9) caregiving (Table 1). It was developed by SHOW scientists in collaboration with 27 faculty and stakeholders spanning different departments at the University of Wisconsin-Madison (See Acknowledgements) and the Wisconsin Department of Health and Family Services.

**Table 1.** SHOW’s COVID-19 Survey Domains.

|  |  |
| --- | --- |
| **COVID-19 – Perceptions, Beliefs, Behaviors** | **Health & Healthcare Access** |
| *History of exposure & testing* | *Chronic health conditions* |
| *Symptoms & hospitalization* | *Flu vaccine history* |
| *Mitigation perceptions & behaviors* | *Medication / treatment access* |
| *Perceived threat* | *Missed appointments* |
| *Coping strategies* | *Delayed procedures/surgeries* |
| *Vaccine willingness\** | *Advance care planning\** |
| *Lasting symptoms\** | *Reproductive health\** |
| **Economic Well Being** | **Mental & Emotional health** |
| *Employment status & job type* | *Anxiety and Stress screener* |
| *Changes in employment* | *Previous Mental Health Diagnosis* |
| *Insurance status & type* | *PTSD screener* |
| *Change in insurance* | *Emotional support* |
| *Retirement funds loss* | *COVID-19 specific stressors* |
| **Food security, diet and housing** | **Information Sources and Literacy** |
| *Use of benefit programs before/after* | *Trusted news sources* |
| *Worried about food lasting* | *Internet access* |
| *Changes in eating habits / meals\** | *Know how to find info on internet\** |
| *Foods consumed more/less\** | **Lifestyle behaviors** |
| *Unable to pay rent/mortgage* | *Sleep quality* |
| *Had to relocate* | *Physical activity* |
| **Personal, social & community context** | *Alcohol consumption* |
| *Resilience* | *Smoking habits* |
| *Social cohesion* | **Caregiving** |
| *Risk Taking, trust, altruism* | *Children /Adults* |
| *Discrimination* | *Stress, strain & coping* |
| *Political Empowerment* | *Family activities* |

\*Topic was added in Wave II and/or Wave III

Where possible, the COVID-19 Survey matched SHOW baseline survey questions to allow for longitudinal comparisons.^29^ Additional survey items were identified using validated instruments and the NIH COVID-19 questions bank^30^ or adapted from other established surveys for comparability across population-based cohorts (Table 2). Ethnicity and race were asked separately and self-identified by the participant at baseline via self-administered questionnaire. SHOW scientists and UW professors developed survey items for topics for which a validated instrument did not exist. Additional survey items were added in Wave II and Wave III, including questions about the impacts of at-home schooling, vaccine hesitancy, long COVID-19 symptoms, and advanced care planning. Surveys and a comparison of survey domains across waves (Supplementary Table 1) are in supplemental materials.

**Table 2.** SHOW COVID-19 survey items and their sources.

| **SHOW COVID-19 Survey Items** | **Source of survey questions** |
| --- | --- |
| COVID-19 testing and symptoms | MESA COVID-19 questionnaire<sup>31</sup> |
| General health status | SF-12<sup>32</sup> |
| Mental Health | Patient Health Questionnaire (PHQ)-4<sup>33</sup> |
| Demographics, smoking, alcohol use, current and past medical conditions | National Health and Nutrition Examination Survey (NHANES)<sup>34</sup> or Behavioral Risk Factor Surveillance System (BRFSS)<sup>35</sup> |
| Healthcare Access | 2018 National Health Interview Survey<sup>36</sup> |
| Reproductive Health | Desire to Avoid Pregnancy<sup>37</sup> |
| Vision and Hearing Impairment | Health and Retirement Survey<sup>38</sup> |
| Food security | USDA food security module and USAR2<sup>39</sup> |

The survey was administered online via Research Electronic Data Capture (REDCap), a secure, web-based software platform for research data capture, hosted at the University of Wisconsin-Madison.^40,41^ Staff entered phone survey data into participant’s unique REDcap survey link. The phone survey was tailored to reduce length and ease administration (response options were removed, collapsed, or made open text for field interview to capture response). Figure 1A depicts the data collection timeline: Wave I occurred May 18, 2020 - July 5, 2020; Wave II February 1 - March 11, 2021; and Wave III from July 8 - August 14, 2021.

Participants received a $25 gift-card for each completed survey wave. The study was approved by the University of Wisconsin-Madison Health Sciences Institutional Review Board. All survey data was downloaded from REDCap and analyzed using SASv.4. Participants were considered rural if their residence was in census block group defined by the U.S. Census Bureau as having fewer than 2500 people^42^ and participant reported household income was used to determine poverty status using the U.S. Federal Poverty Level (FPL) via the U.S. Department of Health and Human Services’ annual poverty guidelines.^43^ Quantitative frequencies and percents were reported as asked in the survey. T-tests and pooled variance were used to compare response rates by group. If cell sizes were less than 10, response options were collapsed and notated in results table.

## RESULTS

### Response rates and characteristics of participants vs. non-participants

A total of n=1,403, n=1,889 and n=1,854 completed the online COVID-19 Survey for waves I, II and III, resulting in response rates of 46%, 63%, and 57%, respectively (Figure 2). Supplementary Table 2 summarizes demographic characteristics of non-participants (n = 3,266) and participants (n = 2,463), using baseline information from individuals most recent (pre-COVID-19) participation in SHOW. Those who participated in at least one wave of the online survey were more likely to be older (mean age 56 years vs. 52 years), female (62% vs. 52%) and non-Hispanic white (89% vs. 81%), have at least a Bachelor’s degree (44% vs. 24%), and live above 200% of the Federal Poverty Level (76% vs. 62%) when compared to those who were invited and did not participate in any wave (Supplementary Table 2). Response rate did not vary significantly by urbanicity or chronic medical condition.

There was a total of n=2,304 unique participants who completed one or more online waves (response rate 71%). A total of n=1,090 participants completed all three waves, whereas n=552 completed only one of the three survey waves. See Figure 3 and Supplementary Table 3 for participation by wave. An additional n=190 completed at least one of the three phone survey waves; n=55, n=122, and n=139 completed waves I, II, and III, respectively.

**Figure 3.**
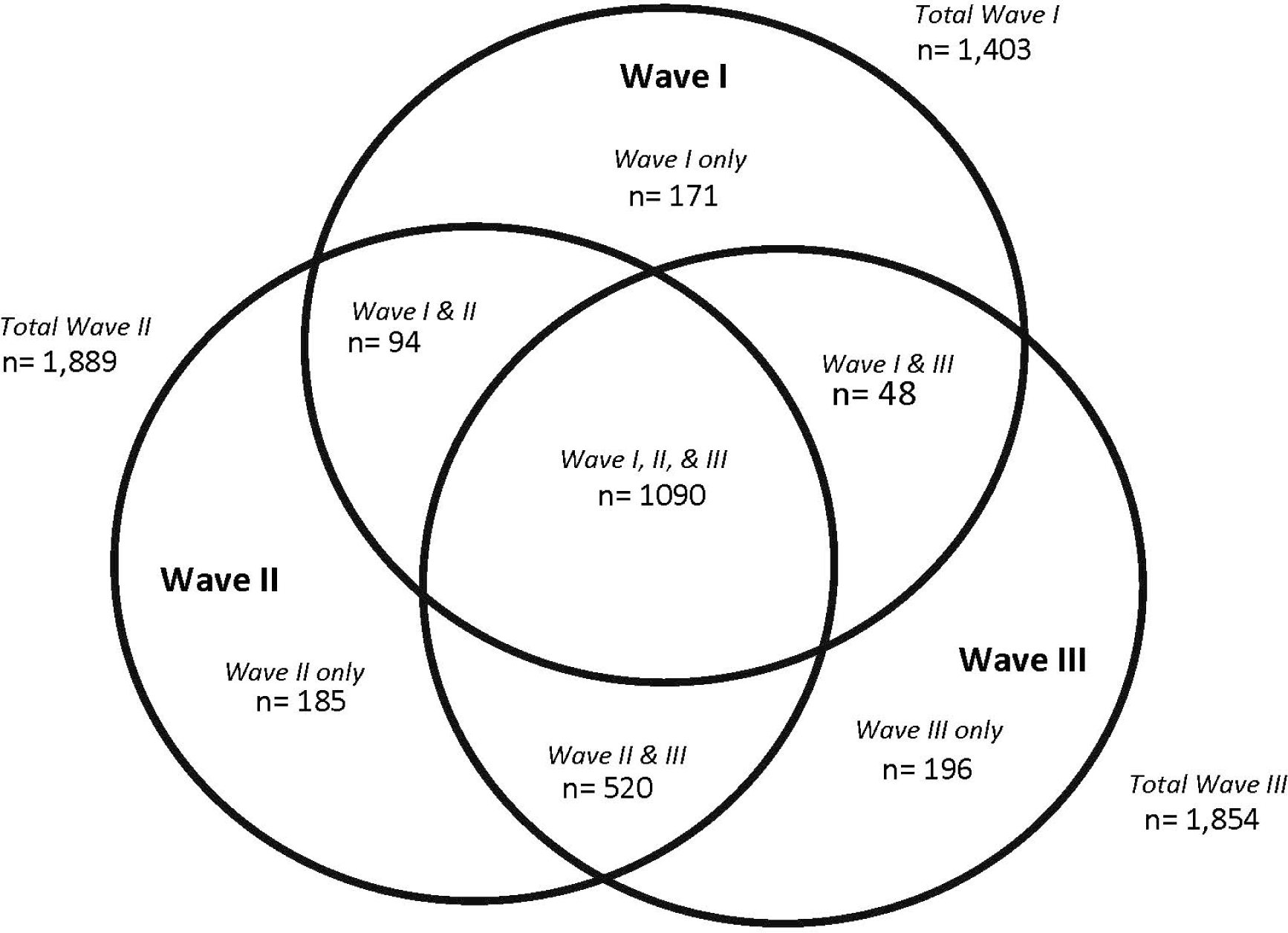
Venn Diagram of participation by online survey waves completed

Table 3 depicts demographics and characteristics of online COVID-19 Survey participants by wave. The distribution of demographics and characteristics were similar across waves. Participants had a mean age of 56 years (SD 15) with majority age 35-79 years (over 85%), just over 60% female, and about 90% identified as White. Approximately 50% had a bachelor’s degree or higher education, and 80% reported a household income greater than 200% of the FPL. One-third of participants lived in a rural area. And 10-30% of participants had at least one medical condition unrelated to COVID-19 (10% reported having cancer, and 30% having high blood pressure).

**Table 3.** Demographic and general characteristics of participants in the online COVID-19 Survey, by wave of data collection, reported as n (%)^a,b^.

|  | **Wave I**<br>May-June<br>2020<br>n = 1403 | **Wave II**<br>Jan-Feb 2021<br>n = 1889 | **Wave III**<br>June-Aug<br>2021<br>n = 1854 |
| --- | --- | --- | --- |
| **Sample Characteristic** |  |  |  |
| Age (years), mean (SD) | 56 (15) | 57 (15) | 57 (15) |
| Age (years) |  |  |  |
| 18 to 34 | 152 (11) | 152 (8) | 173 (9) |
| 35 to 64 | 783 (56) | 1043 (55) | 979 (53) |
| 65 to 79 | 419 (30) | 613 (32) | 626 (34) |
| 80 and older | 49 (3) | 81 (4) | 76 (4) |
| Hispanic or Latino ethnicity<sup>c</sup> |  |  |  |
| No | 1369 (98) | 1839 (97) | 1810 (98) |
| Yes | 34 (2) | 49 (3) | 43 (2) |
| Race, all races identified (not mutually exclusive)<sup>c,d</sup> |  |  |  |
| White | 1275 (91) | 1729 (92) | 1702 (92) |
| Black or African American | 110 (8) | 134 (7) | 124 (7) |
| Asian | 18 (1) | 26 (1) | 28 (2) |
| Native Hawaiian or other Pacific Islander | 5 (0) | 5 (0) | 5 (0) |
| American Indian or Alaska Native | 33 (2) | 51 (3) | 43 (2) |
| Other (written response) | 23 (2) | 29 (2) | 24 (1) |
| Gender<sup>e</sup> |  |  |  |
| Female | 875 (64) | 1135 (61) | 1134 (62) |
| Male | 500 (36) | 725 (39) | 688 (38) |
| Sexual identity |  |  |  |
| Straight | 1311 (96) | 1784 (96) | 1723 (96) |
| Lesbian, gay, bisexual, or other | 50 (4) | 69 (4) | 81 (4) |
| Education |  |  |  |
| HS degree or lower | 230 (17) | 301 (16) | 291 (16) |
| Some college or Associate's degree | 448 (33) | 651 (35) | 642 (35) |
| Bachelor's degree or higher | 698 (51) | 915 (49) | 888 (49) |
| Poverty<sup>f</sup> |  |  |  |
| ≤ 200% FPL | 260 (21) | 330 (20) | 296 (18) |
| > 200% FPL | 999 (79) | 1352 (80) | 1348 (82) |
| Census 2010 urban / rural classification<sup>c,g</sup> |  |  |  |
| Urban | 962 (69) | 1246 (66) | 1238 (67) |
| Rural | 441 (31) | 643 (34) | 616 (33) |
| Current medical conditions unrelated to COVID-19<sup>h</sup> |  |  |  |
| High blood pressure or hypertension | 397 (29) | 600 (32) | 572 (31) |
| High cholesterol or hyperlipidemia | 333 (24) | 471 (25) | 463 (25) |
| Anxiety | 269 (20) | 371 (20) | 358 (20) |
| Depression | 250 (18) | 367 (20) | 346 (19) |
| Asthma | 182 (13) | 212 (11) | 217 (12) |
| Diabetes | 137 (10) | 213 (11) | 210 (11) |
| Cancer | 131 (10) | 179 (10) | 165 (9) |

**Abbreviations:** COPD=chronic obstructive pulmonary disease; COVID-19=coronavirus disease 2019; FPL=federal poverty level; HS=high school; SD=standard deviation; SHOW=Survey of the Health of Wisconsin.
<sup>a</sup>Variable distributions are reported as n (%), unless otherwise specified.
<sup>b</sup>Frequencies may not add to the total sample size due to missing responses.
<sup>c</sup>Ethnicity, race, and urban / rural classification are calculated using the most recent core SHOW data for each individual.
<sup>d</sup>Each race category includes the total number of participants who selected that race in a multiple response question, regardless of other races identified. Participants who wrote in responses to the question on race are included in "Other (written response)", however, their written responses were not recoded into existing categories.
<sup>e</sup>Due to sample size, "trans male" was recoded as "male", "trans-female" was recoded as "female", and "gender non-conforming" was recoded as missing.
<sup>f</sup>Calculations use the annual poverty guidelines from Health and Human Services (HHS) from the year in which the data were collected. Household Income was collected categorically, so the midpoints for the income categories were used in the calculation. The number of people supported by the family income was capped, so responses of "over 7 people" were recoded as 8.
<sup>g</sup>Urban / rural classification is based on residential address. For adult participants who were minors when they most recently participated in core SHOW, the address of the adult who consented to their participation is used.
<sup>h</sup>Participants were asked, "We would like to know about any current medical conditions, not related to COVID-19. Please indicate if you have EVER been told by a doctor or health care professional that you had any of the following. Select all that apply." The wording of response options in the table is true to that of the original question. Participants who selected "Don't know", or "Other medical condition. Please describe:" are also included in the denominator, however, written responses were not recoded into existing categories.

Compared to online survey participants, phone interview participants were older (mean age 71 years compared to 56 years), non-White (30% vs. 10%), high school or lower education (50-60% compared to 16%) and at or below 200% of the FPL (50% vs. 20%) (Supplementary table 4). Phone survey participants were more likely to report having other medical conditions compared to the online participants; 58% vs 29% reported high blood pressure, and 20% vs. 10% reported having cancer, for phone vs. online participants.

### COVID-19 experiences and perceptions during 2020-2021

Changes in COVID-19 experiences and perceptions across COVID-19 Survey domains are summarized in Table 4. Eleven percent (in wave I) thought they may have had COVID-19 since COVID-19 started, which jumped to 22% in wave II and remained at 20% in wave III. Delays in healthcare due to COVID-19 increased from 43% (wave I) to 53% (wave III). Less than 10% of participants reported rationing medication due to the pandemic, which saw a slight increase across the survey waves. Eleven percent and 15% of participants scored as having depression and anxiety, respectively, which stayed consistent in wave I-II and decreased slightly in wave III for both conditions.

**Table 4:** Experiences of participants in the online COVID-19 Survey, by wave of data collection, reported as n (%)^a,b^.

| Sample Characteristic | Wave I<br>May-June 2020<br>n = 1403 | Wave II<br>Jan-Feb 2021<br>n = 1889 | Wave III<br>June-Aug 2021<br>n = 1854 |
| --- | --- | --- | --- |
| **Health and Healthcare Access** |  |  |  |
| Thought may have had COVID-19 since COVID-19 started | 133 (11) | 364 (22) | 337 (20) |
| Delayed care due to COVID-19<sup>c</sup> | 607 (43) | 934 (49) | 983 (53) |
| Rationing of medication<sup>c,d</sup> | 63 (4) | 120 (6) | 135 (7) |
| **Mental and Emotional Health** |  |  |  |
| Depression (PHQ-2) | 145 (11) | 202 (11) | 159 (9) |
| Anxiety (GAD-2) | 207 (15) | 276 (15) | 226 (12) |
| Anxiety and Depression (PHQ-4) |  |  |  |
| Normal (0-2) | 884 (64) | 1181 (64) | 1261 (70) |
| Mild (3-5) | 327 (24) | 430 (23) | 369 (20) |
| Moderate (6-8) | 106 (8) | 154 (8) | 108 (6) |
| Severe (9-12) | 53 (4) | 68 (4) | 63 (4) |
| **Information Sources and Literacy** |  |  |  |
| Trust local public health officials or WI DHS | 1029 (73) | 1303 (69) | 1297 (70) |
| Trust HHS or CDC | 1014 (72) | 1329 (70) | 1281 (69) |
| **Lifestyle Behaviors** |  |  |  |
| Change in alcohol consumption |  |  |  |
| A little to a lot more | 299 (23) | 352 (19) | 293 (16) |
| About the same | 793 (61) | 1203 (64) | 1254 (69) |
| A little lower to much lower | 198 (15) | 313 (17) | 280 (15) |
| Change in physical or usual activities due to COVID-19 |  |  |  |
| A little less to much less active | 605 (44) | 898 (48) | 625 (34) |
| About the same | 458 (33) | 692 (37) | 882 (49) |
| A bit more to much more active | 310 (13) | 271 (15) | 308 (17) |
| **Caregiving** |  |  |  |
| Care for ≥ 1 child in household | 353 (26) | 453 (24) | 356 (23) |
| Care for ≥ 1 adult with illness or disability in household | 71 (5) | 89 (5) | 107 (6) |
| **COVID-19 Beliefs** |  |  |  |
| Believe COVID-19 is a threat in my household |  |  |  |
| Agree to Strongly Agree | 691 (50) | 1053 (56) | 648 (35) |
| Neutral | 230 (16) | 257 (14) | 233 (13) |
| Disagree to Strongly Disagree | 473 (34) | 560 (30) |  |
|  |  |  | 953 (52) |
| Experienced stigma or discrimination due to COVID-19 |  |  |  |
| Yes | 81 (6) | 194 (11) | 199 (11) |
| **Financial** |  |  |  |
| Unable to pay rent/mortgage due to COVID-19 |  |  |  |
| Yes | 185 (13) | 107 (6) | 85 (5) |
| No | 1040 (75) | 1620 (86) | 1634 (89) |
| Not Applicable | 160 (12) | 152 (8) | 116 (6) |
| Concerned about losing job in next 3 month<sup>e</sup> |  |  |  |
| Somewhat to very worried | 82 (12) | 78 (8) | 47 (5) |
| Unsure | 64 (9) | 73 (7) | 48 (5) |
| Not worried or not very worried | 535 (79) | 851 (85) | 854 (90) |

<sup>a</sup>Variable distributions are reported as n (%), unless otherwise specified.
<sup>b</sup>Frequencies may not add to the total sample size due to missing responses.
<sup>c</sup>Time frame for the response changed across waves from Wave 1 (ever) to Wave 2 (since July 1, 2020) to Wave 3 (since February 1, 2021).
<sup>d</sup>Rationing of medication included: taking less medication because you could not get it or could not afford it or delayed filling a prescription due to COVID-19.
<sup>e</sup>Demoninator includes only participants who reported working.

Half the participants agreed or strongly agreed COVID-19 was a threat to their household, which increased to 56% (wave II) and then dropped to 35% (wave III). Six percent reported experiencing stigma or discrimination due to COVID-19, which jumped to 11 percent (waves II-III). Majority reported trusting local public health officials, the state health department, and federal agencies, like the U.S. Center for Disease Control (CDC) (69-73% across all three waves) as sources for information about the pandemic.

A quarter of participants reported caring for at least one child in their household and 5% reported caring for an adult in the home with illness or disability, consistent across survey waves. Thirteen percent reported being unable to pay rent or mortgage due to COVID-19 in wave I, which dropped to 6% and 5% in wave II and wave III; a similar trend was seen with concerns about losing their job in the next 3 months. Yet, food insecurity remained stable throughout the survey waves with 11-13% reporting food insecurity.

### Impact of COVID-19 on the lived experience

Data from this study have been used to identify differences in the lived experience seen across multiple determinants of health. Wave 1 revealed Non-whites were 2-3 times more likely to think they had COVID-19 or have told they have COVID-19 compared to Whites, females were more likely than males to experience a partial or full deduction of paid wages or hours, adults with children in home were more likely to report moderate to high levels of stress compared to adults without children in the home.^28^This study’s data also highlight how mental health problems may be exacerbated by COVID-19 lockdowns and affect those with sensory impairments more severely due to lack of in-person contacts.^44,45^

About a quarter of participants reported caring for at least one child in their household and 5% reported caring for an adult in the home with illness or disability, which stayed consistent across survey waves. Thirteen percent reported being unable to pay rent or mortgage due to COVID-19 in wave I, which dropped to 6% and 5% in wave II and wave II; a similar trend was seen with concerns about losing their job in the next 3 months. Yet, food insecurity remained stable throughout the survey waves with 11-13% reporting being food insecure.

### Impact of COVID-19 on the lived experience

Data from this study have been used to identify differences in the lived experience seen across multiple determinants of health. Wave 1 revealed Non-whites were 2-3 times more likely to think they had COVID-19 or have told they have COVID-19 compared to Whites, females were more likely than males to experience a partial or full deduction of paid wages or hours, adults with children in home were more likely to report moderate to high levels of stress compared to adults without children in the home.^28^This study’s data also highlight how mental health problems may be exacerbated by COVID-19 lockdowns and affect those with sensory impairments more severely due to lack of in-person contacts.^44,45^

### Strengths and Limitations

A primary strength of the SHOW COVID-19 cohort is its extension from a longstanding, well characterized, cohort of Wisconsin residents with pre-COVID-19 longitudinal survey data, physical measurements and stored biological specimens. The SHOW includes over 2,000 health indicators, outcomes, and social determinants of health; residential locations allow for geospatial linkages to contextual and environmental data; and bio-banked specimens (stool and blood derivatives) are available for microbiome, genetic, and epigenetic analyses. The highly engaged cohort enabled a high response rate in this study. The three longitudinal waves of data collection allow for causal analysis. Furthermore, the cohort consented to linkage to public data (vitals, vaccinations, cancer registries, etc.) and consented to be contacted for follow-up studies, enabling continued growth of the infrastructure, data collection, and impactful scientific findings. The COVID-19 Survey was an interdisciplinary effort of a diverse group of expert investigators who came together to develop and launch the survey quickly in May 2020, resulting in data spanning a broad array of health factors and indicators relating to COVID-19.

The pandemic also posed limitations for recruitment and participation. In-person and mail-based recruitment methods were not possible during the early stages of the pandemic. This may have resulted in a lower response rate, especially among older and rural populations. Pre-pandemic recruitment methods included in-person or mailed invites in addition to e-invites or phone recruitment. Yet, we were able to modify the web-based survey for a phone-based interview. This is an important feature of this study, as the phone-survey participants differed significantly from the online survey participants (they were much older, less educated, poorer, and more likely to be non-White). While two different modes of survey administration pose a limitation for data users, ensuring greater representation in the data outweighs the bias and measurement error due to mode and question text changes.

## Conclusions

The SHOW COVID-19 cohort and COVID-19 Survey offers a unique resource for advancing understanding the impact of the COVID-19 pandemic across a diverse population-based sample. Few studies have such a diverse sample that builds on an existing real-world cohort. Data are extensive and offer researchers numerous ongoing opportunities to explore COVID-19’s impact on population health equity, and insight for managing future pandemics. Data from the COVID-19 Survey are currently being linked with COVID-19 serology data to assess objectively how social and behavioral impacts of the pandemic were associated with exposure and contraction of COVID-19. Additional ongoing analyses include examining how COVID-19 impacted longitudinal changes in sleep quantity and quality, and access to healthcare services, treatment, and screenings, and assessing the factors associated with long COVID symptoms. Limited data are free and publicly available, and complete data are available upon request. The cohort is on-going and available for continued follow-up studies.

## ETHICS APPROVAL

The study was approved by the University of Wisconsin-Madison Health Sciences Institutional Review Board [2013-0251-CP132].

## ACKNOWLEDGEMENTS

The authors would like to thank for the SHOW participants for the continued engagement and participation in studies, as well as the University of Wisconsin Survey Center, SHOW administrative, field, and scientific staff for their assistance in study implementation, data collection and data cleaning. Funding for the Survey of the Health of Wisconsin (SHOW) was provided by the Wisconsin Partnership Program PERC Award (233 AAG9971). Survey development was made possible due to the generous contributions from several investigators at the University of Wisconsin: Janean Dilworth-Bart, Kristin Litzelman, Heather Kirkorian, Margaret Kerr, Tiffany Green, Natasha Merten, Marietou Ouayogode, Deborah Ehrenthal, Rebecca Myerson, Lydia Ashton, Thomas Oliver, Maureen Smith, Amy Trentham-Dietz, Maureen Durkin, Jane Mahoney, Marguerite Burns, and Jonathan Patz.

## AUTHOR CONTRIBUTIONS

AAS managed data collection, directed data curation and analysis, and was the primary writer. ENB contributed to the literature review, writing and analysis. SL and JB curated and processed the data and generated tables and figures and analyses used. MN assisted with study design and implementation. MCW and AB assisted to sampling, data, collection and early data curation. LCB, CE, JL, PP, and AS contributed to survey design and survey content. KMCM directed and oversaw and study, led the conceptualization, design, and manuscript objectives.

## FUNDING

This work was supported by the Wisconsin Partnership Program (WPP) PERC Award [233 PRJ 25DJ and WPP4444]. WPP supports efforts to improve the health of the people of Wisconsin by supporting partnerships between the University of Wisconsin School of Medicine and Public Health and local, regional, and statewide groups to address the most important health issues in Wisconsin.

## CONFLICT OF INTEREST

The authors declare that the research was conducted in the absence of any commercial or financial relationships that could be construed as a potential conflict of interest.

## DATA AVAILABILITY & COLLABORATION

The SHOW COVID-19 study investigators encourage use of COVID-19 Survey data and stored biospecimen for future grants and publications. Limited use data from all waves of the COVID-19 Survey is free and publicly available. Access to complete survey data from COVID-19 Survey, and access to baseline SHOW survey data, are subject to fee-for-service charges available for use via a data request process. The SHOW study investigators encourage collaboration and use of the SHOW COVID-19 cohort, data, and stored biospecimen for future grants and publications. Participants in the SHOW COVID-19 cohort have consented to be contacted for future follow-up studies and SHOW study investigators are available to assist researchers interested in following up the cohort and expanding data collection. For those interested in using the data, or leveraging SHOW COVID-19 cohort or SHOW cohort for ancillary studies, contact the corresponding author or visit the SHOW website: https://show.wisc.edu/

## Notes

### Competing Interest Statement

The authors have declared no competing interest.

### Summary of Updates

The following section of text was removed as it was mistakenly included twice: “across all three waves) as sources for information about the pandemic, and a similar percent of participants trusted federal agencies, like the U.S. Center for Disease Control (CDC). About a quarter of participants reported caring for at least one child in their household and 5% reported caring for an adult in the home with illness or disability, which stayed consistent across survey waves. Thirteen percent reported being unable to pay rent or mortgage due to COVID-19 in wave I, which dropped to 6% and 5% in wave II and wave II; a similar trend was seen with concerns about losing their job in the next 3 months. Yet, food insecurity remained stable throughout the survey waves with 11-13% reporting being food insecure.”

