## Supplementary material for "The SHOW COVID-19 cohort: methods and rationale for examining the statewide impact of COVID-19 on the social determinants of health": Supplmentary Table 4

| **Supplementary Table 4:** Demographic and general characteristics of participants in the phone COVID-19 Survey, by wave of data collection, reported as n (%)^a,b^ | | | |
| --- | --- | --- | --- |
| **Sample Characteristic** | **Wave I**  May-June 2020  n = 55 | **Wave II**  Jan-Feb 2021  n = 122 | **Wave III**  June-Aug 2021  n = 139 |
| Age (years), mean (SD) | 71 (10) | 70 (11) | 71 (11) |
| Age (years) |  |  |  |
| 18 to 34 | 0 (0) | 0 (0) | 1 (1) |
| 35 to 64 | 12 (22) | 38 (31) | 34 (24) |
| 65 to 79 | 31 (56) | 54 (44) | 63 (45) |
| 80 and older | 12 (22) | 30 (25) | 41 (30) |
| Race, all races identified (not mutually exclusive)^c,d^ |  |  |  |
| White | 42 (76) | 86 (70) | 102 (73) |
| Black or African American | 12 (22) | 37 (30) | 37 (27) |
| Other | 3 (5) | 9 (7) | 12 (9) |
| Gender^e^ |  |  |  |
| Female | 35 (64) | 77 (63) | 93 (67) |
| Male | 20 (36) | 45 (37) | 45 (33) |
| Education |  |  |  |
| HS degree or lower | 34 (62) | 75 (61) | 73 (53) |
| Some college or Associate's degree | 13 (24) | 35 (29) | 45 (32) |
| Bachelor's degree or higher | 8 (15) | 12 (10) | 21 (15) |
| Poverty^f^ |  |  |  |
| ≤ 200% FPL | 30 (71) | 66 (62) | 63 (52) |
| > 200% FPL | 12 (29) | 40 (38) | 59 (48) |
| Census 2010 urban / rural classification^c,g^ |  |  |  |
| Urban | 27 (49) | 83 (68) | 92 (66) |
| Rural | 28 (51) | 39 (32) | 47 (34) |
| Current medical conditions unrelated to COVID-19 |  |  |  |
| High blood pressure or hypertension | 32 (58) | 84 (73) | 90 (65) |
| High cholesterol or hyperlipidemia | 30 (55) | 76 (66) | 90 (65) |
| Anxiety | 14 (25) | 30 (26) | 39 (28) |
| Depression | 18 (33) | 32 (28) | 40 (29) |
| Asthma | 13 (24) | 30 (26) | 29 (21) |
| Diabetes | 14 (25) | 37 (32) | 43 (31) |
| Cancer | 11 (20) | 24 (21) | 29 (21) |
| **Abbreviations**: COPD=chronic obstructive pulmonary disease; COVID-19=coronavirus disease 2019; FPL=federal poverty level; HS=high school; SD=standard deviation; SHOW=Survey of the Health of Wisconsin.  ^a^Variable distributions are reported as n (%), unless otherwise specified.  ^b^Frequencies may not add to the total sample size due to missing responses.  ^c^Race and urban / rural classification are calculated using the most recent core SHOW data for each individual.  ^d^Each race category includes the total number of participants who selected that race in a multiple response question, regardless of other races identified. Due to sample size, "Other" includes participants who identified as Asian, Native Hawaiian or other Pacific Islander, or American Indian or Alaska Native. Participants who wrote in responses to the question on race are also included in "Other"; their written responses were not recoded into existing categories.  ^e^Due to sample size, "trans male" was recoded as "male", "trans-female" was recoded as "female", and "gender non-conforming" was recoded as missing.  ^f^Calculations use the annual poverty guidelines from Health and Human Services (HHS) from the year in which the data were collected. The number of people supported by the family income was capped, so responses of "over 7 people" were recoded as 8.  ^g^Urban / rural classification is based on residential address.  ^h^Participants were asked, "I am going to read you a list of health conditions. Please tell me "yes or no" if you have EVER been told by a doctor or health care professional that you had that condition." The wording of response options in the table is true to that of the original question. Participants who selected "Don't know", or "Other medical condition. Please describe:" are also included in the denominator, however, written responses were not recoded into existing categories. | | | |
