## Supplementary material for "The SHOW COVID-19 cohort: methods and rationale for examining the statewide impact of COVID-19 on the social determinants of health": Supplmentary Table 1

| **Supplementary Table 1:** SHOW COVID-19 Survey domains across waves of data collection | | | |
| --- | --- | --- | --- |
| **Survey Domains** | **Wave I**  May-June 2020  n = 1403 | **Wave II**  Jan-Feb 2021  n = 1889 | **Wave III**  June-Aug 2021  n = 1854 |
| COVID-19 – Perceptions, Beliefs, Behaviors |  |  |  |
| *History of exposure & testing* | x | x* | x* |
| *Symptoms & hospitalization* | x | x* | x* |
| *Mitigation perceptions & behaviors* | x | x | x |
| *Perceived threat* | x | x | x |
| *Perceived effectiveness, safety of mitigation behaviors* | x* | x | x* |
| *Coping strategies* | x | x | x |
| *Vaccine history* | -- | -- | x |
| *Vaccine willingness* | -- | x | *--* |
| *Lasting symptoms* | -- | -- | x |
| *Difficulty communication due to mask* | -- | -- | x |
| Economic Well Being |  |  |  |
| *Employment status & job type* | x | x | x |
| *Changes in employment* | x | x | x |
| *Insurance status & type* | x | -- | -- |
| *Change in insurance* | x | -- | x |
| *Retirement funds loss* | -- | x | x |
| *Financial concerns* | x | x* | x* |
| Food, housing, and transportation |  |  |  |
| *Use of benefit programs before CV19* | x | x | x |
| *Use of benefit programs during, due to CV19* | x* | x | x |
| *Worried about food lasting* | x | x | x |
| *Received food, necessities from family or friends* | x | -- | -- |
| *Changes in eating habits / meals* | -- | x | x |
| *Foods consumed more/less* | -- | x | x |
| *Unable to pay rent/mortgage* | x | x | x |
| *Had to relocate* | x | x | x |
| *Worried forced to move* | -- | -- | x |
| *Use of public transportation* | x | x | x |
| Personal, social & community context |  |  |  |
| *Resilience* | x | x* | x* |
| *Social cohesion* | x | x | x |
| *Risk Taking, trust, altruism* | -- | x | x* |
| *Stigma or discrimination* | x | x | x |
| *Political Empowerment* | -- | x | -- |
| *Community and neighborhood* | x | -- | x |
| Health & Healthcare Access |  |  |  |
| *Positive or suspected positive CV19 test* | x | x | x |
| *Chronic health conditions* | x | x | x |
| *Current treatment of health conditions* | x | -- | -- |
| *Flu vaccine history* | -- | x | -- |
| *Medication / treatment access* | x | x | x |
| *Missed or postponed appointments* | x | x | x |
| *Delayed procedures/surgeries* | x | x | x |
| *Unmet healthcare needs* | x | x | x |
| *Advance care planning* | -- | x | x |
| *Reproductive health* | -- | x | x |
| *Hearing, vision rating* | x | x | x* |
| *Sought advice from healthcare professional* | x | -- | -- |
| Mental & Emotional health |  |  |  |
| *Anxiety and Stress screener* | x | x | x |
| *Previous Mental Health Diagnosis* | x | x | x |
| *Utilization of mental health services* | x | x | x |
| *PTSD screener* | -- | -- | x |
| *Emotional support* | -- | x | x |
| *COVID-19 specific stressors* | x | x* | x* |
| *Depression and loneliness* | x | x | x |
| *Stressful events or situations* | -- | -- | x |
| Information Sources and Literacy |  |  |  |
| *Trusted news sources* | x | x | x |
| *Internet access* | x | -- | -- |
| *Know how to find info on internet* | -- | x | x |
| Lifestyle behaviors |  |  |  |
| *Sleep quality* | x | x | x |
| *Physical activity* | x | x | x |
| *Usual activities* | x | x | x |
| *Alcohol consumption* | x* | x | x |
| *Smoking habits* | x | x | x |
| Caregiving |  |  |  |
| *Children /Adults* | x | x | x |
| *Stress, strain & coping* | x | x | x |
| *Parenting sentiment* | -- | x | x |
| *Family activities* | x | x | -- |
| *Health of children, families* | x | -- | -- |
| -- domain not represented within wave  *Moderate to major changes to questions within domain, but domain still represented within that wave  **Note**: There are slight changes to wording across waves in regard to time windows (i.e. Wave 1 for the most part does not have specific time windows, while W2/W3 reference specific time windows); these are not denoted in this table | | | |
