## Supplementary material for "The SHOW COVID-19 cohort: methods and rationale for examining the statewide impact of COVID-19 on the social determinants of health": Supplmentary Table 2

| **Supplementary Table 2:** Comparison of baseline demographic and general characteristics^a^ of eligible non-participants and participants^b^ in the COVID-19 Survey, reported as n (%)^c,d^ | | | |
| --- | --- | --- | --- |
| **Sample Characteristic** | **Non-Participants**  n = 3266 | **Participants**  n = 2463 | ***P* Value** |
| Age (years), mean (SD)^e^ | 52 (18) | 56 (15) | < 0.0001 |
| Age (years)^e^ |  |  |  |
| 18 to 34 | 590 (18) | 252 (10) |  |
| 35 to 64 | 1744 (54) | 1345 (55) |  |
| 65 to 79 | 706 (22) | 753 (31) |  |
| 80 and older | 200 (6) | 109 (4) | < 0.0001 |
| Hispanic or Latino ethnicity |  |  |  |
| No | 3098 (95) | 2397 (97) | < 0.0001 |
| Yes | 158 (5) | 64 (3) |  |
| Race, all races identified (not mutually exclusive)^f^ |  |  |  |
| White | 2639 (81) | 2187 (89) | < 0.0001 |
| Black or African American | 516 (16) | 241 (10) | < 0.0001 |
| Asian | 42 (1) | 33 (1) | 0.9 |
| Native Hawaiian or other Pacific Islander | 14 (0) | 7 (0) | 0.4 |
| American Indian or Alaska Native | 176 (5) | 72 (3) | < 0.0001 |
| Other (written response) | 125 (4) | 41 (2) | < 0.0001 |
| Gender |  |  |  |
| Female | 1698 (52) | 1518 (62) | < 0.0001 |
| Male | 1568 (48) | 945 (38) |  |
| Census 2010 urban / rural classification^g^ |  |  |  |
| Urban | 2208 (68) | 1645 (67) | 0.4 |
| Rural | 1043 (32) | 818 (33) |  |
| Education^h^ |  |  |  |
| HS degree or lower | 1140 (37) | 500 (21) | < 0.0001 |
| Some college or Associate's degree | 1189 (39) | 874 (36) |  |
| Bachelor's degree or higher | 746 (24) | 1060 (44) |  |
| Poverty^i^ |  |  |  |
| ≤ 200% FPL | 1177 (38) | 566 (24) | < 0.0001 |
| > 200% FPL | 1900 (62) | 1825 (76) |  |
| Medical conditions^h^ |  |  |  |
| High blood pressure or hypertension | 1053 (34) | 767 (32) | 0.03 |
| High cholesterol or hyperlipidemia | 964 (32) | 839 (35) | 0.02 |
| Anxiety^j^ | 481 (17) | 402 (17) | 0.5 |
| Depression^j^ | 613 (22) | 538 (22) | 0.8 |
| Asthma | 544 (18) | 379 (16) | 0.04 |
| Diabetes | 392 (13) | 258 (11) | 0.01 |
| Cancer^j^ | 264 (10) | 321 (13) | < 0.0001 |
| ^a^Baseline characteristics are calculated using the most recent core SHOW data for each individual.  ^b^Eligible non-participants are individuals eligible for at least one wave of the COVID-19 impact survey. Participants are individuals who participated in at least one wave of the phone or online survey.  ^c^Variable distributions are reported as n (%), unless otherwise specified.  ^d^Frequencies may not add to the total sample size due to missing responses.  ^e^Age as of 01 May 2020. Some participants were under 18 years of age as of this date but were at least 18 years old when they partook in the COVID-19 impact survey.  ^f^Each race category includes the total number of participants who selected that race in a multiple response question, regardless of other races identified. Participants who wrote in responses to the question on race are included in "Other (written response)", however, their written responses were not recoded into existing categories.  ^g^Urban / rural classification is based on residential address. For some participants who were minors when they most recently partook in core SHOW, the address of the adult who consented to their participation is used.  ^h^Education and medical conditions are only reported for individuals aged 18 years or older when they most recently partook in core SHOW.  ^i^Calculations use the annual poverty guidelines from U.S. Health and Human Services (HHS) from the year in which the data were collected. Household Income was collected categorically, so the midpoints for the income categories were used in the calculation. For some participants who were minors when they most recently partook in core SHOW, the poverty status of the adult who consented to their participation is used.  ^j^Participants were asked if they had ever been told by a doctor or health care professional that they had that medical condition. Anxiety, depression, and cancer were asked at a later time point for 2008-2013 participants than for 2014-2019 participants. Diabetes excludes participants who only had diabetes during pregnancy.  **Abbreviations**: COVID-19=coronavirus disease 2019; df=degrees of freedom; FPL=federal poverty level; HS=high school; SD=standard deviation; SHOW=Survey of the Health of Wisconsin. | | | |
