## Supplementary material for "The SHOW COVID-19 cohort: methods and rationale for examining the statewide impact of COVID-19 on the social determinants of health": Supplmentary Table 3

| **Supplementary Table 3**. Participation in online SHOW COVID-19 Survey by survey waves completed | | |
| --- | --- | --- |
| Waves completed: | N | % |
| Wave I only | 171 | 7.4 |
| Wave II only | 185 | 8.0 |
| Wave III only | 196 | 8.5 |
| Wave I & II | 94 | 4.1 |
| Wave I & III | 48 | 2.1 |
| Wave II & III | 520 | 22.6 |
| Wave I, II, & III | 1090 | 47.3 |
| Total number of unique individuals: | 2,304 | 100 |
